# Systolic Blood Pressure and Survival to Very Old Age. Results from the Women’s Health Initiative

**DOI:** 10.1101/2023.06.22.23291783

**Authors:** Bernhard Haring, Chris A. Andrews, Kathleen Hovey, Aladdin H. Shadyab, Andrea LaCroix, Lisa Warsinger Martin, Milagros C. Rosal, Lewis H. Kuller, Elena Salmoirago-Blotcher, Nazmus Saquib, Patrick Koo, Deepika Laddu, Marcia L. Stefanick, JoAnn E. Manson, Sylvia Wassertheil-Smoller, Michael J. LaMonte

## Abstract

**Background:** The association between systolic blood pressure (SBP) and longevity is not fully understood. We aimed to determine survival probabilities to age 90 for various SBP levels among women aged ≥ 65 years with or without BP medication.

**Methods:** We analyzed blood pressure data from participants in the Women’s Health Initiative (n=16,570) who were aged 65 or older and without history of cardiovascular disease, diabetes or cancer. Blood pressure was measured at baseline (1993-1998) and then annually through 2005. The outcome was defined as survival to age 90 with follow-up until February 28, 2020.

**Results:** During a follow-up of 18 years, 9,723 (59%) of 16,570 women survived to age 90. The SBP associated with the highest probability of survival was about 120mmHg regardless of age. Compared to an SBP between 110 and 130 mmHg, women with uncontrolled SBP had a lower survival probability across all age groups and with or without BP medication. A 65-year-old women on BP medication with an interpolated SBP between 110 and 130 mmHg in 80% of the first 5 years of follow-up had a 31% (95% confidence interval, 24%, 38%) absolute survival probability. For those with 20% time in range, the probability was 21% (95% confidence interval, 16%, 26%).

**Conclusions:** An SBP level below 130 mmHg was found to be associated with longevity among older women. The longer SBP was controlled at a level between 110 and 130 mmHg, the higher the survival probability to age 90. Preventing age-related rises in SBP and increasing the time with controlled BP levels constitute important measures for achieving longevity.

**Clinical Perspective:** *What is new ?:*

- The age-related rise in SBP is commonly regarded as inevitable and intensified SBP treatment in older adults is still controversial, as strict BP control in older adults has been related to a higher risk of mortality.
- Based on a real-world national cohort, an SBP level below 130 mmHg was associated with the highest survival probability to age 90 years in women with or without BP medication.
- The longer the time women had their SBP controlled at ≥ 110 and <130mmHg, the higher the probability of survival to age 90.

*What are the clinical implications ?:*

- The age-related BP estimates in conjunction with survival probabilities to age 90 presented clearly emphasize the importance of maintaining well-controlled BP levels even at older age.
- Preventive measures and risk factor control to ensure a constant relatively low SBP pattern during ageing are warranted.

## Introduction

A healthy lifestyle has been associated with an increased life expectancy.^1^ Cardiovascular risk factors such as smoking, insufficient physical activity, diabetes, hypercholesterolemia or an unhealthy diet have been shown to reduce longevity.^2–6^ However, while some studies have characterized SBP over a wide range of ages, and have examined associations of SBP with subclinical and clinical cardiovascular endpoints, the pattern of SBP associated with survival to age ≥90 years remains unclear.^7–14^ As the population is rapidly ageing with adults and particularly women greater than 65 years representing the fastest growing segment of the U.S. adult population, a better understanding of which SBP levels are associated with survival to very old age is important to identify strategies to lengthen lifespan and healthspan.^15, 16^

In this analysis using BP measurements from a large U.S. cohort, we sought to elucidate which SBP levels in women older than 65 years and free of major chronic diseases were associated with the highest probability of surviving to age 90. We hypothesized that in women with or without BP medication, adhering to recommended SBP target ranges (i.e., SBP < 130 mmHg) would be associated with the highest probability of survival to age 90.

## Methods

### Study Population

The study population consisted of women enrolled in the randomized clinical trials of the Women’s Health Initiative. Details of the Women’s Health Initiative Clinical Trials have been described previously.^17, 18^ In brief, participating women were recruited between 1993 and 1998 (baseline) at 40 clinical centers in the United States, were generally healthy and postmenopausal. After enrollment, they had their BP measured annually as part of their routine clinical examinations (n=68,132). All participants provided written informed consent, and institutional review board approval was received by all participating institutions.

For the present analysis, women aged ≥ 65 years and who were eligible to survive to age 90 (i.e., who were or would have turned 90 on or before February 28, 2020) as of the latest outcome assessment (N=23,643) and who had a baseline BP measurement and at least 5 of the first 7 annual BP measurements (N=20,146) were included. For reasons of competing risk that could affect likelihood of survival, we excluded women with a history of cancer (except nonmelanoma skin cancer) (N=436), history of cardiovascular disease (coronary heart disease [myocardial infarction, coronary artery revascularization procedure, unstable angina], stroke or heart failure) (N=1,281), history of diabetes (N=1,298) or missing survival status (N=1,384). Our final study population consisted of 16,570 women who were followed-up until February 28, 2020, for a mean of 18 years.

### Assessment of Blood Pressure

BP was measured at baseline (1993-1998) and at each annual study visit with the use of a standardized measurement protocol by WHI staff trained in taking BP in clinical settings.^17–20^ Participants were asked to sit with both feet flat on the floor and to rest without talking for five minutes before measuring her BP. Appropriate cuff bladder size was determined at each visit based on arm circumference. BP was measured in the right arm with a stethoscope and mercury sphygmomanometer; 2 measures, taken 30 seconds apart, were recorded. The BP at each visit was defined as the average of the 2 readings. Up to 7 BP measurements per woman were included in the present study (Supplemental Figure 1).

### Time in Target Range

Time in target range (TTR) expresses the proportion of time recorded SBP measurements were within a certain window.^21^ Linear interpolation between SBP measurements was used to estimate TTR during the five years after WHI enrollment. The recommended target range was defined as SBP ≥110 and <130 mmHg following prior analyses.^22^ The upper limit of 130 mmHg was specifically selected to match current guideline-recommended SBP target levels in older persons.^23^

### Survival

Participants were classified as having survived to very old age (defined to be 90yrs) or to have died before this age. Trained physician adjudicators confirmed deaths according to hospital records, autopsy or coroner’s reports, or death certificates. Periodic linkage to the National Death Index was performed for all participants, including those lost to follow-up, for verification if medical records or death certificates were not available. All outcomes were centrally adjudicated.^17, 19, 24, 25^

### Covariates

Participants completed questionnaires assessing age, race and ethnicity, education, annual income, marital status, diet quality (Healthy Eating Index (HEI)), smoking status, alcohol intake, hormone therapy use and recreational physical activity at baseline.^26^ Trained clinic staff measured height and weight using a calibrated clinical scale and stadiometer. Body mass index (BMI) was calculated as weight in kilograms divided by height in meters squared. Antihypertensive medication data were collected during in-person clinic visits at baseline.^26^

### Statistical Analysis

Descriptive statistics were computed for baseline demographic variables in the overall cohort eligible to survive to age 90, according to SBP categories at baseline, and according to survival status at the end of follow-up. Continuous variables were summarized by mean and standard deviations; categorical variables by frequencies and percentages. The absolute probability of surviving to age 90 years was estimated for all combinations of SBP and age at measurement using a generalized additive logistic regression model with a tensor product smooth of age at visit and SBP. The SBP that maximized survival was estimated for each age and a 95% confidence interval was generated from a standard error computed from 1000 bootstrap replications (resampled at participant level). Unadjusted models are presented to describe general associations of survival with SBP and age. Heatmaps display the survival probability for combinations of SBP (between 80 and 200 mmHg) and age at measurement (between 65 and 82 years) for (a) the whole study sample, women (b) with and (c) without BP medication at baseline. In addition to the figures, survival probabilities (with 95% bootstrap confidence intervals) were tabulated for selected SBP values and ages. In supplementary analyses, we repeated these calculations for various DBP values (between 55 and 100 mmHg) and age combinations. Ratios of survival probabilities (and CIs) between SBP levels at fixed ages were estimated. As a sensitivity analysis to account for possible confounding, we expanded our aforementioned models using SBP and age to include main effects for race and ethnicity, education, annual income, marital status, smoking status, alcohol consumption, body mass index, recreational physical activity, diet quality (Healthy Eating Index), hormone therapy use and history of hypertension. Finally, unadjusted absolute survival probabilities were estimated as a function of baseline age and the percentage of time in the SBP target range (SBP ≥ 110 and <130mmHg) over the next five years for the whole study sample and separately for women with and without BP medication at baseline. As a sensitivity analysis we adjusted our models with the same covariates described above. Analyses were performed using SAS software (v 9.4, SAS Inc, Cary, NC) and with the mgcv package in the R statistical computing environment (v 4.2.1, R Foundation for Statistical Computing, Vienna, Austria).^27^

### Data Availability

The data, analytic methods, and study materials are made available to other researchers for purposes of reproducing the results or replicating the procedure. The data underlying our work can be obtained through 2 mechanisms. First, interested investigators can contact the Women’s Health Initiative Coordinating Center. Details about the procedures for data request can be found online (www.whi.org). Second, most data from the Women’s Health Initiative can be also be obtained from BioLINCC, a repository maintained by the National Heart, Lung, and Blood Institute. The BioLINCC website includes detailed information about the available data and the process to obtain such data (https://biolincc.nhlbi.nih.gov/home/).

## Results

During a mean follow-up of 18 years among 16,570 women, 42% survived to age 90 and 58% died before age 90. Women had a mean age of 70.6 years (standard deviation 3.4 years, range 55 to 79) at baseline and 75% of the study sample had 7 SBP measurements between the ages of 60 and 82.

Participants’ characteristics overall and by measured SBP categories at baseline are presented in Table 1. Women at recommended SBP levels (i.e., < 130 mmHg) were slightly younger, more likely to be non-Hispanic, white and married, to have higher education and income, to have a normal BMI, a non-smoking history and to be more physically active and a current alcohol consumer. Use of BP medication was less prevalent and these women had a slightly healthier diet. Overall, the lower the SBP level, the lower the observed DBP level.

**Table 1:** Baseline characteristics by baseline Systolic Blood Pressure (mmHg)

| Characteristic | Overall<br>(N= 16,570) | <110<br>(N= 1,327) | ≥ 110 and<br><130<br>(N= 6,134) | 130-139<br>(N= 3,700) | 140-159<br>(N= 4,089) | ≥160<br>(N=1,320) |
| --- | --- | --- | --- | --- | --- | --- |
| **Age at baseline (Mean (SD))** | 70.6 (3.4) | 70.1 (3.4) | 70.3 (3.4) | 70.6 (3.4) | 70.9 (3.4) | 71.5 (3.5) |
| **Race and Ethnicity** |  |  |  |  |  |  |
| Non-Hispanic Asian | 263 (1.6) | 16 (1.2) | 88 (1.4) | 45 (1.2) | 86 (2.1) | 28 (2.1) |
| Non-Hispanic Black | 967 (5.8) | 40 (3.0) | 289 (4.7) | 224 (6.1) | 301 (7.4) | 113 (8.6) |
| Non-Hispanic White | 14,732 (88.9) | 1,223 (92.2) | 5,548 (90.4) | 3,300 (89.2) | 3,552 (86.9) | 1,109 (84.0) |
| Hispanic | 346 (2.1) | 28 (2.1) | 121 (2.0) | 71 (1.9) | 87 (2.1) | 39 (3.0) |
| Unknown/not listed above | 262 (1.6) | 20 (1.5) | 88 (1.4) | 60 (1.6) | 63 (1.5) | 31 (2.3) |
| **Education** |  |  |  |  |  |  |
| High school or less | 4,225 (25.6) | 281 (21.3) | 1,510 (24.7) | 981 (26.7) | 1,082 (26.6) | 371 (28.4) |
| Some college or vocational training | 6,566 (39.8) | 457 (34.6) | 2,422 (39.6) | 1,469 (39.9) | 1,657 (40.7) | 561 (43.0) |
| College graduate | 5,701 (34.6) | 583 (44.1) | 2,181 (35.7) | 1,231 (33.4) | 1,332 (32.7) | 374 (28.6) |
| **Income** |  |  |  |  |  |  |
| <20,000 US\$ | 3,610 (22.6) | 239 (18.7) | 1,267 (21.4) | 797 (22.4) | 947 (24.1) | 360 (27.9) |
| 20,000-<50,000 US\$ | 8,315 (52.1) | 659 (51.6) | 3,079 (52.1) | 1,863 (52.4) | 2,076 (52.8) | 638 (49.5) |
| ≥50,000 US\$ | 4,036 (25.3) | 380 (29.7) | 1,563 (26.5) | 895 (25.2) | 906 (23.1) | 292 (22.6) |
| **Marital status** |  |  |  |  |  |  |
| Married/living as married | 9,249 (56.0) | 746 (56.4) | 3,429 (56.1) | 2,096 (56.9) | 2,252 (55.3) | 726 (55.2) |
| Widowed | 4,891 (29.6) | 375 (28.4) | 1,765 (28.9) | 1,083 (29.4) | 1,258 (30.9) | 410 (31.2) |
| Divorced/separated | 1,749 (10.6) | 149 (11.3) | 671 (11.0) | 385 (10.4) | 403 (9.9) | 141 (10.7) |
| Never married | 619 (3.7) | 52 (3.9) | 247 (4.0) | 121 (3.3) | 161 (4.0) | 38 (2.9) |
| **Smoking status** |  |  |  |  |  |  |
| Never Smoked | 9,086 (55.5) | 657 (50.0) | 3,388 (55.9) | 2,045 (55.9) | 2,255 (56.0) | 741 (56.8) |
| Past Smoker | 6,468 (39.5) | 548 (41.7) | 2,358 (38.9) | 1,464 (40.0) | 1,599 (39.7) | 499 (38.3) |
| Current Smoker | 811 (5.0) | 108 (8.2) | 320 (5.3) | 147 (4.0) | 172 (4.3) | 64 (4.9) |
| **Alcohol intake** |  |  |  |  |  |  |
| Nondrinker | 1,873 (11.4) | 121 (9.2) | 652 (10.7) | 413 (11.3) | 503 (12.4) | 184 (14.0) |
| Past drinker | 2,786 (16.9) | 223 (16.9) | 994 (16.3) | 628 (17.1) | 701 (17.3) | 240 (18.3) |
| Current drinker | 11,780 (71.7) | 976 (73.9) | 4,450 (73.0) | 2,629 (71.6) | 2,837 (70.2) | 888 (67.7) |
| **Body Mass Index (kg/m<sup>2</sup>)** |  |  |  |  |  |  |
| Normal (<25) | 5,118 (31.0) | 646 (49.0) | 2,024 (33.1) | 982 (26.7) | 1,084 (26.6) | 382 (29.0) |
| Overweight (25-<30) | 6,364 (38.6) | 445 (33.8) | 2,417 (39.6) | 1,435 (39.0) | 1,559 (38.3) | 508 (38.6) |
| Obese (≥30) | 5,012 (30.4) | 227 (17.2) | 1,666 (27.3) | 1,266 (34.4) | 1,428 (35.1) | 425 (32.3) |
| **Recreational physical activity**<br>**(MET hours/week) (Mean (SD))** | 11.6 (12.7) | 13.6 (14.2) | 11.7 (12.7) | 11.2 (12.0) | 11.1 (12.7) | 11.0 (12.4) |
| **HT use** |  |  |  |  |  |  |
| Never | 9,083 (54.8) | 733 (55.3) | 3,366 (54.9) | 2,036 (55.1) | 2,230 (54.6) | 718 (54.4) |
| Ever | 7,478 (45.2) | 593 (44.7) | 2,765 (45.1) | 1,662 (44.9) | 1,856 (45.4) | 602 (45.6) |
| **HEI (Mean (SD))** | 64.5 (9.8) | 65.2 (9.5) | 64.8 (9.8) | 64.2 (9.9) | 64.1 (9.7) | 64.5 (10.1) |
| **Blood pressure medication use** | 5,755 (34.7) | 205 (15.4) | 1,646 (26.8) | 1,368 (37.0) | 1,811 (44.3) | 725 (54.9) |
| **DBP (mmHg) (Mean (SD))** | 74.6 (9.3) | 64.7 (6.8) | 71.3 (7.4) | 75.8 (7.7) | 79.0 (8.6) | 83.4 (9.0) |

Data presented as N (%) unless otherwise noted
Abbreviations: HT – Hormone Therapy, HEI – Healthy eating index, HTN – hypertension, SBP – systolic blood pressure, DBP – diastolic blood pressure

Baseline characteristics overall and by survival to 90 are presented in Supplemental Table 1. Those women who survived to age 90 were slightly older at enrollment (mean age 71.2 vs 69.8 years), more likely to be white of non-Hispanic origin, married, to be a college graduate, and have a higher income. They were also more likely to be overweight/obese, never smokers, current alcohol drinkers, more physically active, and to have a healthier diet. Conversely, women surviving to age 90 were less likely to have a history of hypertension (49% vs 57%) and use BP medication (31% vs 40%) at baseline compared to those not surviving.

Figure 1 shows a heatmap of our main analysis depicting the SBP associated with the highest probability of survival to age 90 across various ages. The overall pattern indicates that, at each age, the probability of surviving to age 90 is highest near 120 mmHg and is lower for greater SBP. An SBP below 130 mmHg was found to be associated with the highest probability of survival to age 90 across all attained age groups (Supplemental Table 2). Women with an SBP between 110 and 130 mmHg at attained ages of 65, 70, 75, and 80 had 37% (95% confidence interval (CI): 34%,42%), 54% (52%, 55%), 65% (64%, 66%) and 76% (74%, 77%) absolute probability to survive to age 90, respectively. When we compared various SBP levels to an SBP between 110 and 130 mmHg, the survival probability tended to be lower across all age strata with higher SBP levels (Table 2). In sensitivity analysis, when we adjusted our main analysis for socio-demographic and lifestyle factors, the SBP pattern associated with surviving to age 90 again continued to show consistency across ages with an SBP below 130mmHg to be associated with the highest probability to achieve longevity (Supplemental Figure 2 and Supplemental Table 3). We stratified our results by baseline BP medication use (Figure 2 a/b) and found the same pattern with slightly higher survival probability among women not on BP medication (Supplemental Table 4) and lower among women on BP medication (Supplemental Table 5).

**Figure 1.**
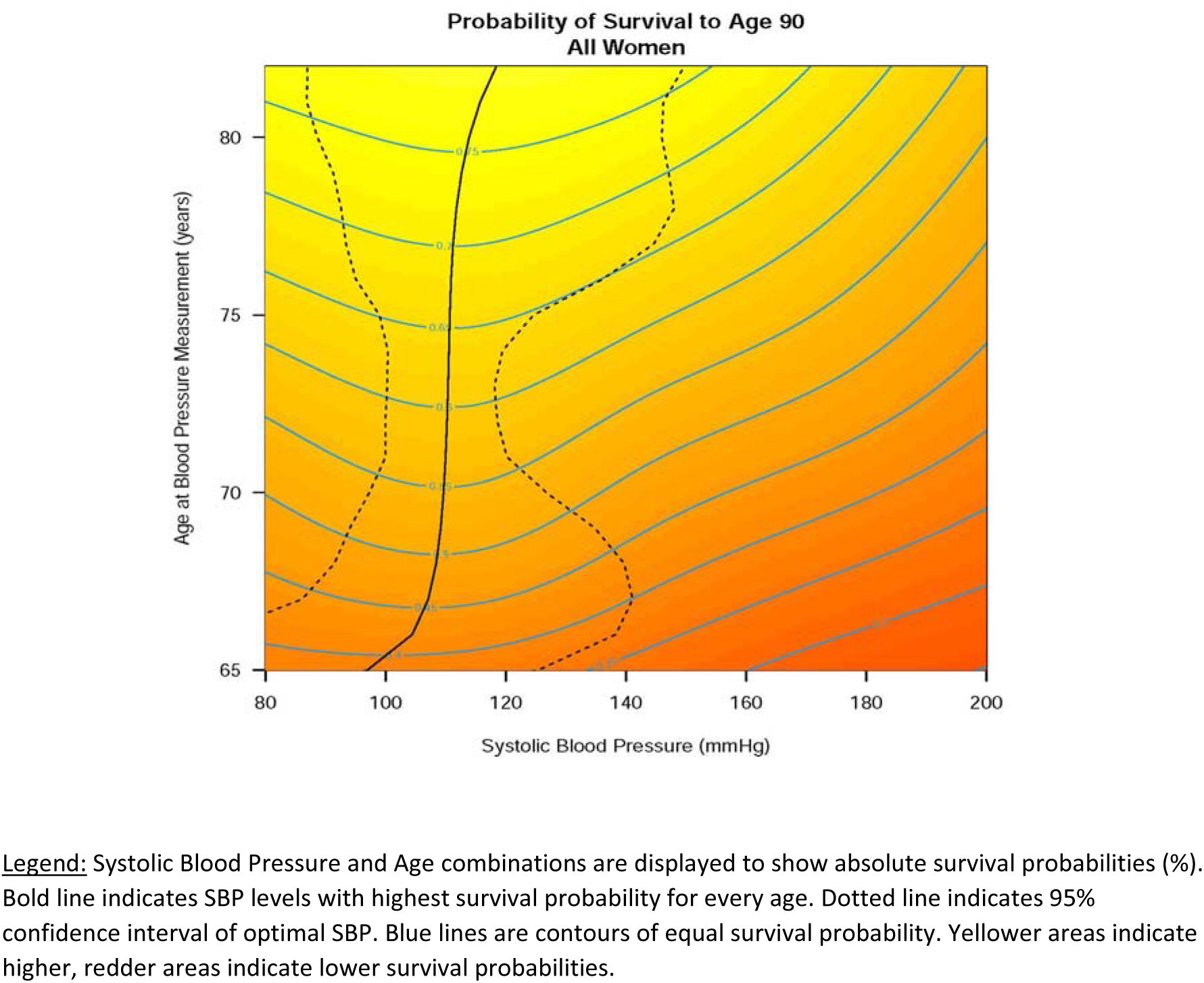
Survival Probability to age 90 years by Age and Systolic Blood Pressure.

**Figure 2a.**
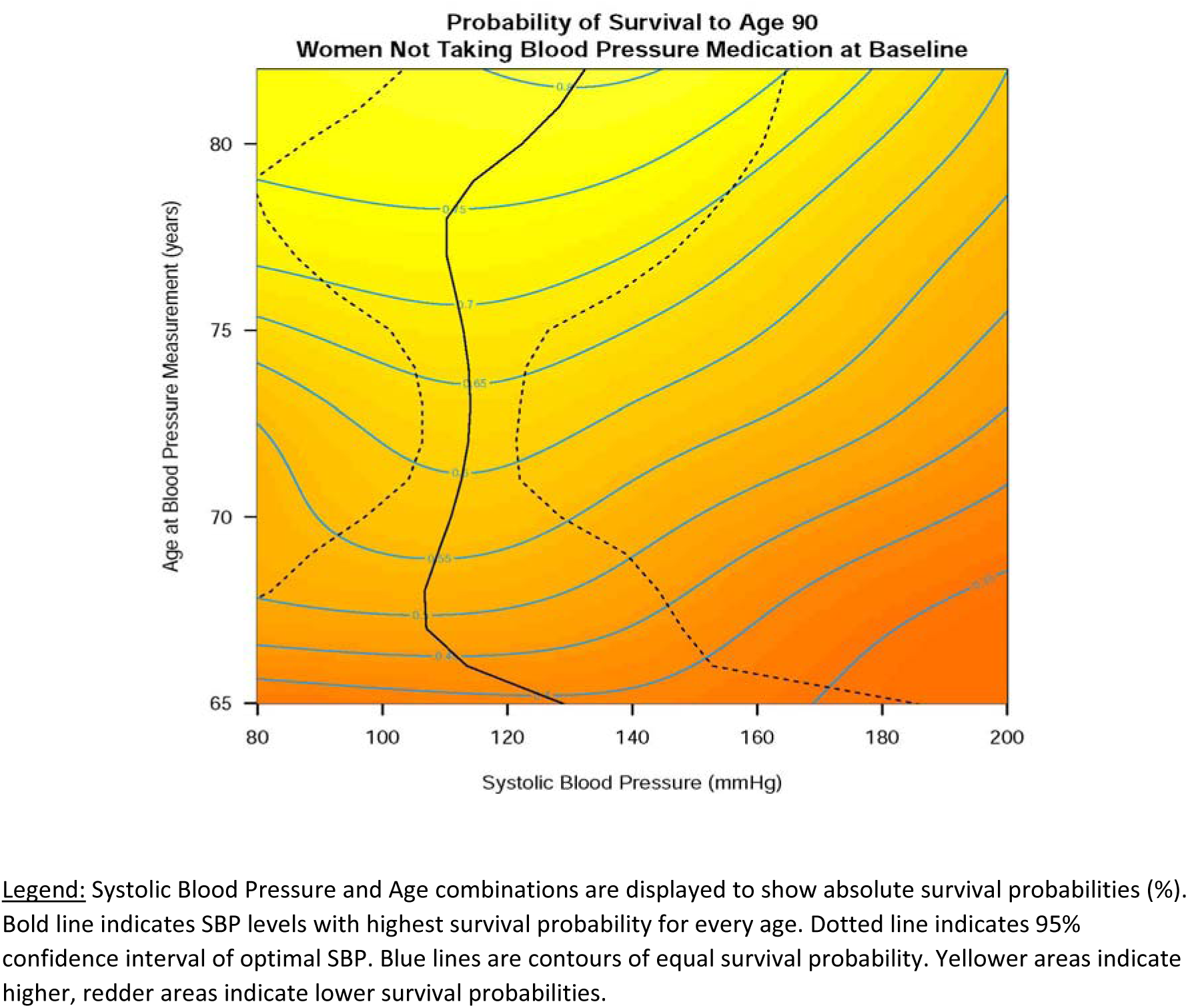
Survival Probability to age 90 by Age and Systolic Blood Pressure among women not on blood pressure medication.

**Figure 2b.**
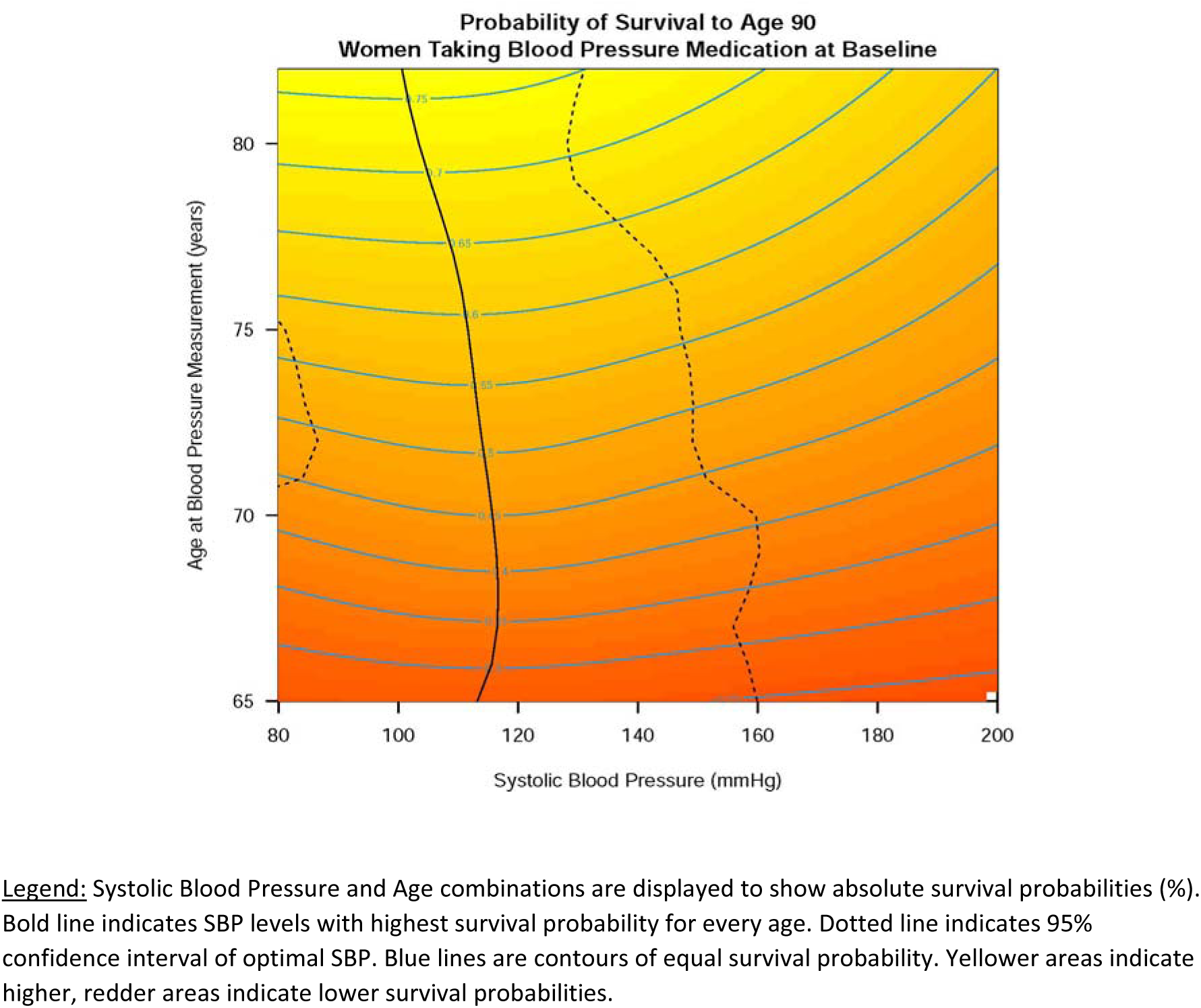
Survival Probability to age 90 by Age and Systolic Blood Pressure among women on blood pressure medication.

**Table 2.** Survival Probability to age 90 by Age and Systolic Blood Pressure among all women and by blood pressure medication status (risk ratio, reference SBP ≥ 110 and <130 mmHg)

| Age<br>(years) | Systolic Blood Pressure (mmHg) |  |  |  |  |
| --- | --- | --- | --- | --- | --- |
| | <110 | $\geq 110$ and <130 | 130-139 | 140-159 | $\geq 160$ |
| **Overall (N=16,570)** |  |  |  |  |  |
| **80** | 1.00 (0.98,1.03) | 1 (ref) | 0.98 (0.96,1.00) | 0.95 (0.92,0.98) | 0.89 (0.85,0.94) |
| **75** | 1.00 (0.99,1.03) | 1 (ref) | 0.96 (0.95,0.98) | 0.91 (0.89,0.94) | 0.86 (0.83,0.90) |
| **70** | 1.01 (0.98,1.04) | 1 (ref) | 0.93 (0.92,0.96) | 0.86 (0.83,0.90) | 0.81 (0.76,0.85) |
| **65** | 1.03 (0.94,1.24) | 1 (ref) | 0.93 (0.83,1.00) | 0.86 (0.74,0.99) | 0.79 (0.63,1.15) |
| **Not on BP medication (N=10,815)** |  |  |  |  |  |
| **80** | 0.99 (0.97,1.03) | 1 (ref) | 0.99 (0.96,1.01) | 0.96 (0.92,0.99) | 0.90 (0.85,0.97) |
| **75** | 1.00 (0.98,1.02) | 1 (ref) | 0.97 (0.95,0.99) | 0.92 (0.89,0.95) | 0.87 (0.82,0.91) |
| **70** | 1.00 (0.97,1.04) | 1 (ref) | 0.95 (0.92,0.97) | 0.88 (0.84,0.92) | 0.81 (0.75,0.87) |
| **65** | 0.98 (0.87,1.14) | 1 (ref) | 1.00 (0.85,1.09) | 0.97 (0.80,1.15) | 0.91 (0.70,1.40) |
| **On BP medication (N=5,755)** |  |  |  |  |  |
| **80** | 1.01 (0.98,1.09) | 1 (ref) | 0.98 (0.96,1.03) | 0.95 (0.92,1.03) | 0.91 (0.85,0.98) |
| **75** | 1.00 (0.96,1.05) | 1 (ref) | 0.98 (0.95,1.01) | 0.95 (0.91,0.99) | 0.91 (0.86,0.97) |
| **70** | 0.99 (0.93,1.10) | 1 (ref) | 0.97 (0.94,1.03) | 0.93 (0.87,1.02) | 0.89 (0.80,0.97) |
| **65** | 1.00 (0.73,2.28) | 1 (ref) | 0.98 (0.81,1.31) | 0.95 (0.72,1.64) | 0.92 (0.68,1.85) |

Survival probabilities by time in target range of SBP across all BP measurements are displayed in Table 3. Women whose SBP was at a target between 110 and 130 mmHg in 80% of the first 5 years of follow-up 40% (36%, 43%), 64% (62%, 66%), 75% (72%, 77%) and 79% (72%, 86%) at age 65, 70, 75, and 80 to survive to age 90 years, respectively, whereas women whose BP was only controlled 20% of the time had a 32% (29%, 35%), 56% (55%, 57%), 68% (67%, 70%) and 82% (77%, 87%) probability at age 65, 70, 75, and 80 of surviving to a very old age. In sensitivity analysis, when we adjusted our main analysis for important covariates, we observed a similar survival pattern (Supplemental Table 6).

**Table 3.** Time in Therapeutic Range of SBP (≥ 110 and <130 mmHg) and Survival Probability to age 90 years by age and blood pressure medication status (%, 95% confidence interval)

| Age (years) | Time in Therapeutic Range of SBP (%) |  |  |  |  |  |
| --- | --- | --- | --- | --- | --- | --- |
|  | 0 | 20 | 40 | 60 | 80 | 100 |
| **Overall (N=16,570)** |  |  |  |  |  |  |
| **80** | 83 (77,89) | 82 (77,87) | 81 (77,85) | 80 (75,85) | 79 (72,86) | 78 (68,87) |
| **75** | 66 (64,69) | 68 (67,70) | 71 (69,72) | 73 (71,74) | 75 (72,77) | 77 (74,79) |
| **70** | 53 (51,55) | 56 (55,57) | 59 (58,60) | 61 (60,63) | 64 (62,66) | 67 (64,69) |
| **65** | 30 (26,34) | 32 (29,35) | 35 (32,37) | 37 (34,40) | 40 (36,43) | 43 (38,48) |
| **Not on BP medication (N=10,815)** |  |  |  |  |  |  |
| **80** | 84 (78,90) | 83 (79,88) | 83 (79,87) | 82 (77,87) | 81 (75,88) | 81 (72,90) |
| **75** | 71 (68,74) | 72 (70,74) | 74 (72,75) | 75 (73,77) | 76 (74,79) | 78 (75,81) |
| **70** | 57 (55,60) | 60 (58,61) | 62 (61,63) | 64 (63,65) | 66 (64,68) | 68 (66,71) |
| **65** | 37 (32,42) | 38 (35,42) | 40 (37,43) | 42 (39,45) | 43 (39,47) | 45 (40,50) |
| **On BP medication (N=5,755)** |  |  |  |  |  |  |
| **80** | 79 (69,89) | 78 (70,86) | 77 (70,85) | 77 (68,86) | 76 (63,89) | 75 (58,92) |
| **75** | 62 (57,66) | 64 (61,67) | 66 (64,69) | 69 (65,72) | 71 (66,75) | 73 (67,78) |
| **70** | 48 (45,51) | 50 (48,53) | 53 (51,55) | 55 (53,58) | 57 (54,61) | 60 (55,64) |
| **65** | 18 (13,24) | 21 (16,26) | 24 (20,28) | 28 (23,32) | 31 (24,38) | 35 (25,45) |

Supplemental Figure 3 shows a heatmap of our main analysis depicting the DBP associated with the highest probability of survival to age 90 across various ages. Overall, the optimal DBP increases until approximately 75 years, while after age 75 years optimal DBP decreases slightly. A DBP between 70 to 80 mmHg was found to be associated with the highest probability of survival to age 90 across all attained age groups (Supplemental Table 7). Women with DBP between 70 and 80 mmHg at attained ages of 65, 70, 75, and 80 had a 36% (95% confidence interval (CI): 32%, 39%), 53% (51%, 54%), 64% (63%, 66%) and 75% (73%, 76%) absolute probability to survive to age 90.

## Discussion

In a large population of women older than 65 years, an SBP level below 130 mmHg was associated with the highest survival probability to age 90 years in women with or without BP medication. The longer the time women maintained their SBP controlled between 110 and 130 mmHg, the higher the survival probability to age 90 years.

There is ample evidence available showing a linear relationship of SBP with cardiovascular disease that extends from high SBP levels (180 mmHg) to relatively low values (110 to 115 mmHg) for SBP.^28–31^ Findings from SPRINT (Systolic Blood Pressure Intervention Trial) demonstrated that intensive pharmacologic therapy to achieve an SBP target of <120 mmHg resulted in fewer cardiovascular events and deaths than did treating to a conventional SBP target of <140 mmHg.^32^ This finding also pertained to ambulatory adults aged 75 years or older.^33^ Most recently, the STEP (Strategy of Blood Pressure Intervention in the Elderly Hypertensive Patients) study provided additional evidence that intensive BP treatment (SBP target, 110 mmHg to <130 mmHg) benefited older hypertensive patients, aged 60 to 80 years, with reduced incidence of cardiovascular events compared to a standard treatment target of 130 mmHg to <150 mmHg.^34^ Yet, this intensified SBP treatment target in older adults is still controversial, as strict BP control in older adults has been related to a higher risk of hypotension, syncope, greater medication burden and falls leading to serious fractures, and mortality.^35–38^ Recent evidence also indicates that, in older patients with limited life expectancy, the harms of intensive BP control sometimes outweigh the benefits as it may require treating 100 older patients (≥60 years) with hypertension for approximately 3 years to prevent 1 major adverse cardiac event.^39^

Our results extend current literature and have noteworthy implications. First, our data based on a real-world national cohort reinforce current BP guidelines and BP target recommendations for older adults aiming at a SBP <130 mmHg, which to this point are based to a large extent on clinical trials.^40^ Similarly, we could also show that the BP target for DBP in older adults should range between 70 and 80 mmHg again largely consistent with current guideline recommendations.^41^ Second, our findings emphasize the significance of BP control and the need for a constant relatively low SBP pattern during ageing to achieve longevity. These data build upon and complement prior results from the Framingham Heart Study in which, a resting SBP that chronically exceeded approximately 120 to 125 mmHg tended to rise at a more rapid rate and signaled incipient hypertension.^42, 43^ Additionally, our DBP results suggest that in adults achieving longevity DBP levels start to gradually decrease only after approximately age 75 reflecting increased vascular stiffness or vascular age at that point of life. This finding nicely extends prior results from the Framingham Heart Study where DBP levels already started to decrease after age 55.^43, 44^ Of note, whereas in all industrialized societies, SBP rises with age, it is not true for some non-industrialized hunter-gatherer societies living a subsistence lifestyle suggesting that an increase in SBP with age is not a fundamental feature of human ageing but highly dependent on lifestyle.^45, 46–48^ Thus, the rise in SBP at later life is neither physiological nor inevitable, which emphasizes the need for preventive strategies. Third, our data offer a glimmer of perspective even for women who are already on BP medication at age 65. When SBP and DBP remained well controlled, we found a respectable probability of reaching a very old age. These findings are in line with results from the Blood Pressure Lowering Treatment Trialists’ Collaboration showing that BP medication should be viewed as an effective tool for preventing cardiovascular disease when an individual’s cardiovascular risk is elevated.^49^

Beyond achieved mean SBP level, time in SBP target range has been recently shown to be another important measure of BP control, as it may provide a more holistic view of an individual patient’s BP control and has been associated with cardiovascular outcomes.^21, 22, 50, 51^ We thus proceeded to reexamine our cohort using ‘time in target range of SBP’ as another parameter in relation to survival probability.^22^ Interestingly, these results emphasize two other key aspects of BP control. We observed that older women whose SBP was between 110 and 130 mmHg across most measurements had a similar or even higher absolute survival probability compared to those not maintaining their SBP in this range. The younger women were in the range of age studied here, the more significant tight BP control became with respect to survival which was particularly apparent in women who were already on BP medication. This adds to the notion that the cumulative exposure to non-controlled SBP levels has long-term harmful effects even after age 65, which again supports tight SBP targets as part of prevention strategies.

The underlying pathophysiological explanation for our findings can likely be derived from databases such as the US National Health and Nutrition Examination Survey (NHANES). In NHANES, among US adults ≥65 years, a normal BP was associated with adherence to a healthy lifestyle and a lower prevalence of adverse health characteristics.^52^ This can also be seen in long-time population based studies such as the Framingham Heart Study, where a higher vascular risk factor burden (i.e. greater BMI, presence of diabetes, smoking, and hyperlipidemia) was associated with higher BP levels over the life course.^9^ The baseline characteristics of our study population reflect this observation. We chose to exclude women with a history of a severe illness a priori since it would necessarily reduce the likelihood of surviving into older age and any treatment for a major chronic disease would be most likely closely intertwined with BP levels. Nonetheless, when we adjusted our analyses for other present socio-demographic and lifestyle risk factors, our main results, indicating a stable SBP pattern with an SBP below 130 mmHg to be associated with longevity, persisted. Moreover, our stratified analysis by BP medication use reveal that, even in the case of an already present cardiovascular risk factor, longevity can be achieved by adhering to currently recommended SBP targets.^23^

Strengths of our analysis include the use of a large well-characterized population-based national cohort of older women with long-term follow-up. The results are based on one of the largest datasets in the world including more than 9,700 women who survived to age 90. We leveraged a large number of available BP measurements conducted over the course of WHI to create models that related SBP to reaching a very old age. BP was assessed using the same standardized procedures for all study visits, which minimizes the risk for measurement error. We created a novel representation of these associations by using ‘heat maps’ that allow for a qualitative visual assessment of the SBP pattern associated with the highest probability of survival to age 90. On the other hand, generalizability is limited, and selection bias is present as our study population consisted of relatively healthy older women who already survived to at least age 65. We cannot extrapolate our findings to male individuals. Moreover, we do not have adequate data on the use of various BP medication classes over time. Prevalence of chronic disease was determined at baseline and not updated during the follow-up because we think that incident chronic disease could be attributed to SBP (e.g. reverse causality) as the population ages. Finally, as our findings are based on an observational cohort, causal inferences cannot be made.

In conclusion, in older women tight BP control with an SBP below 130 mmHg was associated with longevity. The age-related BP estimates in conjunction with survival probabilities to age 90 illustrated here clearly emphasize the importance of maintaining well-controlled BP levels even at older age. These findings should encourage primary care physicians, policy makers, and other stakeholders to ensure adequate BP control in this vulnerable patient group.

## Data Availability

The data, analytic methods, and study materials are made available to other researchers for purposes of reproducing the results or replicating the procedure. The data underlying our work can be obtained through 2 mechanisms. First, interested investigators can contact the Women?s Health Initiative Coordinating Center. Details about the procedures for data request can be found online (www.whi.org). Second, most data from the Women?s Health Initiative can be also be obtained from BioLINCC, a repository maintained by the National Heart, Lung, and Blood Institute. The BioLINCC website includes detailed information about the available data and the process to obtain such data (https://biolincc.nhlbi.nih.gov/home/).

## Acknowledgments

The authors thank the WHI participants, clinical sites, investigators, and staff for their dedicated efforts. A list of current WHI investigators is available online at: https://www-whi-org.s3.us-west-2.amazonaws.com/wp-content/uploads/WHI-Investigator-Short-List.pdf

## Sources of Funding

The WHI program is funded by the National Heart, Lung, and Blood Institute, National Institutes of Health, U.S. Department of Health and Human Services through 75N92021D00001, 75N92021D00002, 75N92021D00003, 75N92021D00004, 75N92021D00005. The opinions expressed in this manuscript are those of the authors and do not necessarily reflect the views of the Department of Health and Human Services/National Institutes of Health. The WHI PAGE 2 study was supported by grant U01HG007376.

## Disclosures

None

## Notes

### Competing Interest Statement

The authors have declared no competing interest.

### Author Declarations

All participants provided written informed consent, and institutional review board approval was received by all participating institutions.

